# Viral Co-infection in COVID-19: Prevalence and Clinical Associations of Human Pegivirus

**DOI:** 10.64898/2026.02.06.26344215

**Authors:** Mathieu Garand, Ching Ching Zhang, Grace Guo, Pranav Kirti, Jack T. Stapleton, Pirooz Eghtesady

## Abstract

**Objective:** This study investigates the prevalence of human pegivirus (HPgV) in SARS-CoV-2-positive patients within the context of viral co-infections that may modulate COVID-19 outcomes and assesses whether HPgV co-infection is associated with COVID-19 severity. HPgV is a widely circulating but rarely monitored human virus with documented immunomodulatory effects in other viral infections, including HIV and Ebola. While HPgV prevalence is low in the general U.S. population (1–2%), it rises markedly in the setting of chronic viral co-infections, particularly HIV (15–40%). Given its immunologic effects and persistence, HPgV represents a biologically plausible but unexplored viral co-factor SARS-CoV-2 infection.

**Methods:** We analyzed four cohorts: SARS-CoV-2–positive individuals, ICU patients with respiratory symptoms but SARS-CoV-2–negative, HIV-positive individuals as a positive control for HPgV detection, and uninfected controls.

**Results:** HPgV prevalence in COVID-19 patients was low (2.1%) and comparable to population estimates. As expected, HPgV prevalence was substantially higher in the HIV cohort (34%), validating assay performance and cohort stratification. Among HPgV-positive COVID-19 cases, most experienced mild disease, with directional trends toward reduced severity despite high baseline risk factors. Healthcare workers in the control group showed unexpectedly elevated HPgV prevalence (9.6%).

**Conclusions:** HPgV is an unmonitored but widely circulating viral co-infection in humans that may influence host responses to SARS-CoV-2. Although limited by small numbers, our findings support further investigation of HPgV and other immunomodulatory viral co-infections in COVID-19. This study suggests that HPgV co-infection may influence COVID-19 outcomes, warranting further investigation.

**Highlights:**

- Systematic screening of human pegivirus (HPgV), an unmonitored viral co-infection, in COVID-19 (n = 634).
- HPgV prevalence in COVID-19 mirrored population estimates but was markedly enriched in HIV (positive control).
- HPgV-positive COVID-19 cases showed trends toward milder clinical outcomes.
- Findings highlight the potential relevance of immunomodulatory viral co-infections in SARS-CoV-2 infection.

**Executive Summary:** This study evaluated the prevalence of human pegivirus (HPgV) co-infection among SARS-CoV-2–positive patients as part of a broader effort to understand how concurrent viral infections may influence COVID-19 severity. HPgV is a largely unmonitored, persistent human virus with well-described immunomodulatory effects in other viral infections, yet it has not been systematically evaluated in COVID-19. We therefore screened HPgV prevalence across COVID-19 cases, comparator cohorts, and an HIV-positive cohort as a positive control due to the well-established high prevalence of HPgV in HIV infection. Our findings indicate that HPgV prevalence in SARS-CoV-2–positive and –negative hospitalized individuals are consistent with the general U.S. population range (approximately 1–5%). Healthcare professionals exhibited a higher HPgV prevalence (∼10%), suggesting that repeated occupational viral exposures may influence infection rates. While limited by small numbers, HPgV co-infection in COVID-19 cases was associated with directional trends toward reduced disease severity, warranting further longitudinal and mechanistic investigation.

**Editor’s summary:** This study identifies human pegivirus as a widely circulating, unmonitored viral co-infection in COVID-19 with potential relevance to disease severity.

## Introduction

The COVID-19 pandemic, caused by SARS-CoV-2, has posed substantial global health challenges, with growing recognition that viral co-infections can shape disease severity, immune responses, and clinical outcomes. Co-infections with other respiratory pathogens such as influenza and RSV have been shown to influence COVID-19 severity (1,2).

Human pegivirus (HPgV), formerly known as GB virus C (GBV-C), is a positive-sense single-stranded RNA virus classified under the Pegivirus genus of the Flaviviridae family (3). Although HPgV is widespread and persistent, it is not routinely monitored in clinical practice. HPgV is typically detected in 1-5% of the U.S. population but reaches prevalence as high as 40% in individuals with HIV. HPgV has also been found co-infecting with several other viruses including Hepatitis B (HBV), CMV, Dengue and Zika, EBV, Ebola and HTLV 1 & 2.

HPgV is known for its persistence (hence, the “pe” or persistent naming in Pegi) in the human host, with viremia observed for years in both immunocompetent and immunocompromised individuals. In multiple viral disease contexts, HPgV has been associated with altered immune activation and improved outcomes, including reduced HIV progression, enhanced vaccine responses, and increased survival in Ebola infection (reviewed in (4)). For example, HPgV is known to interfere with HIV progression, enhance malaria vaccine protection, and improve survival in Ebola patients. Notably, HPgV has shown a protective effect in HIV-infected individuals, reducing disease progression and improving clinical outcomes, including CD4+ T cell counts, HIV RNA viral load, and mortality rates. These protective effects are thought to arise from HPgV-mediated modulation of host immune pathways and HPgV’s interaction with the HIV gp120-gp41 protein complex.

In contrast, HPgV has also been linked to adverse outcomes in specific settings, including an increased risk of non-Hodgkin lymphoma (5–8), underscoring that its immunologic effects are context dependent.

Despite extensive investigation of HIV and other viral diseases, HPgV has not been evaluated in SARS-CoV-2 infection. Structural and immunologic parallels between HIV and SARS-CoV-2 – including similarities between HIV gp41 and the SARS-CoV-2 S2 domain, and documented antibody cross-reactivity – raise the possibility that HPgV-mediated immune modulation could influence COVID-19 outcomes (9–12). While recent studies have explored viral co-infections in large COVID-19 cohorts (1,2), these investigations were limited to a select group of viruses, excluding HPgV. Given that the gp120-gp41 protein complex is a common target for both HPgV and SARS-CoV-2, we posit that HPgV may also affect SARS-CoV-2 pathology (**Figure 1A**). We therefore sought to determine the prevalence of HPgV in COVID-19 and to explore whether this unmonitored viral co-infection is associated with disease severity..

**Figure 1.**
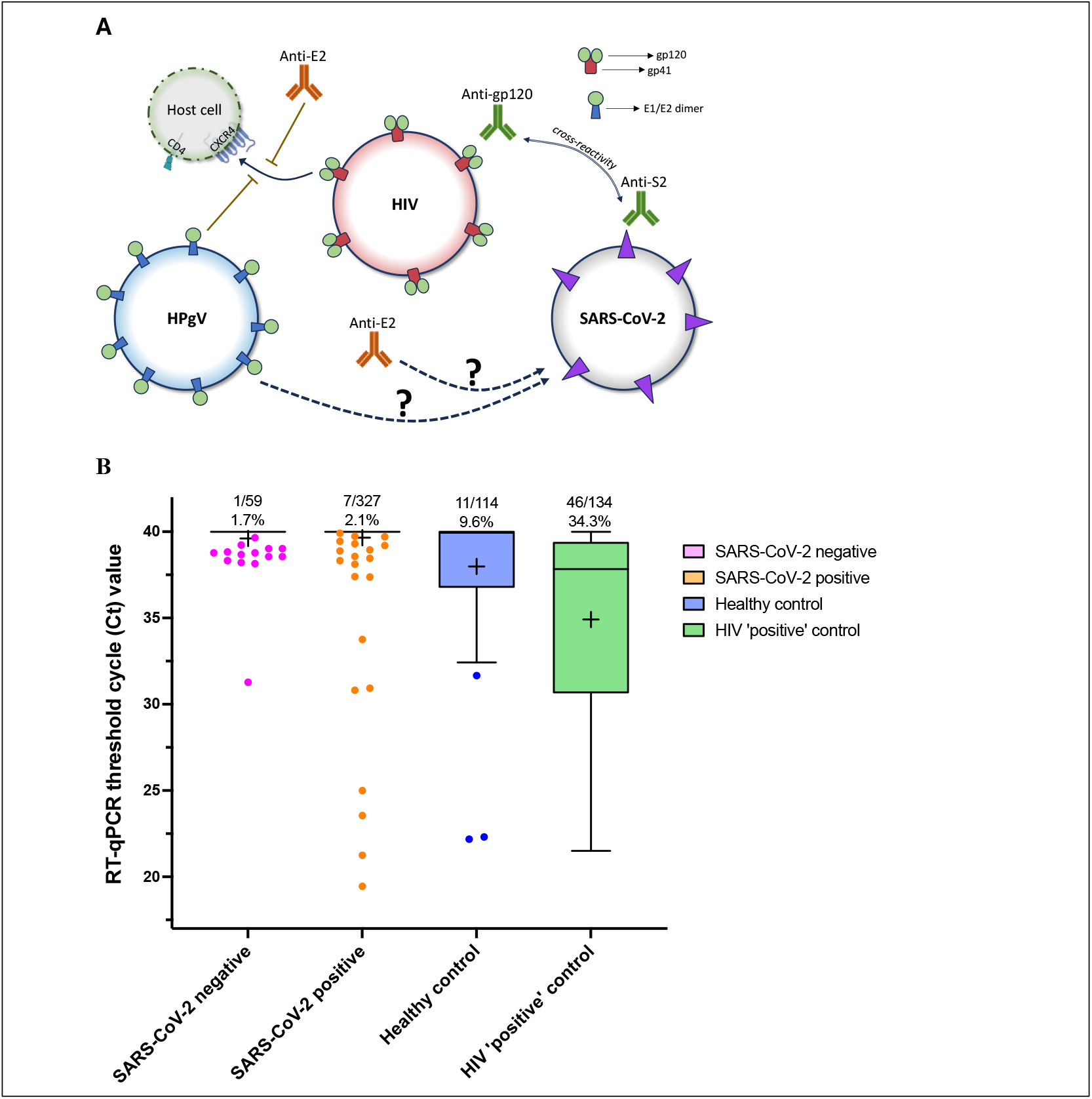
**A)** The interaction between HPgV and HIV is one of the most studied aspects of HPgV’s effect. Research has shown that co-infection of HPgV and HIV results in a protective effect that reduces chances of reactivation of latent HIV. This effect is mediated by HPgV’s interaction with the HIV gp120-gp41 protein complex. Although distinct in genomic organization, HIV’s gp41 and gp120 protein shares remarkable similarities with the spike protein S2. Recent studies have explored viral co-infections in large COVID-19 cohorts, but these studies excluded HPgV. **B)** We performed RT-qPCR to detect HPgV viremia in 634 plasma samples. We assessed the prevalence of HPgV across the four cohorts: COVID-19-negative individuals with respiratory symptoms (n = 59), COVID-19-positive individuals (n = 327), healthy controls (n = 114), and HIV-positive individuals (n = 134). The top and bottom edges of the box represent the upper and lower quartiles (75th and 25th percentiles) respectively. The horizontal line in the box represents the median. The mean indicated as “+”. The whiskers extend 1.5 times the IQR to the upper quartile (for the upper whisker) and the lower quartile (for the lower whisker). Data points within the whisker’s range are not shown - for the SARS-CoV-2 negative and positive groups, many samples had undetectable signal and were assigned the value ‘40’.

## Methods

To investigate HPgV prevalence and clinical associations in COVID-19, we conducted a cross-sectional study using plasma samples collected from 634 participants divided into four cohorts: 327 COVID-19-(RNA)-positive individuals, 59 ICU patients with respiratory symptoms but negative for SARS-CoV-2, 134 HIV-positive patients, and 114 uninfected controls. We included an HIV-positive cohort as a biological positive control, given the well-established high prevalence and persistence of HPgV in HIV infection, allowing validation of HPgV detection and contextual interpretation of prevalence across cohorts. Plasma samples were sourced from Washington University School of Medicine’s COVID-19 Biorepository (14), which was established in 2020 with support from the Institute of Clinical and Translational Sciences (ICTS), The Foundation for Barnes-Jewish Hospital, and the Siteman Cancer Center, with input from the Community Advisory Board of the Institute for Public Health. Clinical specimens were collected during periods of active community transmission, prior to COVID-19 vaccine availability, and were not inactivated. The biorepository was supported by NIH– National Center for Advancing Translational Sciences grant UL1TR002345. The Tissue Procurement Core provided infrastructure for sample processing, storage, and distribution, and coordinated with laboratory personnel across departments to manage the high volume of samples received during the pandemic. The COVID-19-positive and ICU COVID-negative samples were obtained from Washington University’s COVID-19 Biorepository, de-identified, and accompanied by detailed clinical and laboratory data to assess disease severity. Samples from HIV-positive individuals and uninfected controls were obtained from the University of Iowa Virology Clinic and recruited from the Iowa City, IA community. Ethical approval for this study was granted by the Washington University Institutional Review Board (IRB #202210110) and the University of Iowa Institutional Review Board (IRB #201609707).

HPgV RNA detection utilized a TaqMan probe-based one-step RT-qPCR assay following Invitrogen’s SuperScript III Platinum protocol. Total nucleic acids were extracted from 100 μL of plasma using the Zymo Quick DNA/RNA Viral Kit, and HPgV RNA quantification was performed on a QuantStudio3 instrument. We used specific primers and probes targeting HPgV’s 5’UTR region as previously described (15) (see **Supplemental Information** for additional details). A Ct value of less than 35 is classified as HPgV positive, while a Ct value greater than 35 is considered HPgV negative. We repeated the assay for samples falling within 1 Ct of the cut-off and for every positive to confirm the HPgV status.

## Results

Among the COVID-19 cohort of 327 subjects, the prevalence of HPgV co-infection was 2.1% (n = 7), comparable to that found in the general U.S. population (1-5%). COVID-19-negative ICU patients showed a slightly lower prevalence at 1.7%. The HIV-positive cohort exhibited a high HPgV prevalence (34%), consistent with prior literature and confirming the expected enrichment of HPgV in chronic viral infection (reviewed in (4)). Notably, within the uninfected control group of 114 subjects, HPgV prevalence reached 9.6% (n = 11) (**Figure 1B**). This control group was composed primarily of healthcare workers, originally recruited for convenience. While this raises the possibility of cohort-specific exposure patterns, the lack of data on transmission routes and other demographic variables limits further interpretation. Summary of the molecular assay and baseline characteristics of the study cohorts are described in **Table 1 A-B**.

**Table 1.** Descriptive cohort table.

| RT-qPCR summary |  |  |  |
| --- | --- | --- | --- |
| Threshold Cycle for HPgV RT-qPCR | Positive: Ct < 35 | Negative: Ct > 35 | Sum |
| HC, n = 114 (%) | 11 (9.65%) | 103 (90.35%) | 114 |
| HIV, n = 134 (%) | 46 (34.33%) | 88 (65.67%) | 134 |
| Covid-Pos, n = 327 (%) | 7 (2.14%) | 320 (97.86%) | 327 |
| Covid-Neg, n = 59 (%) | 1 (1.69%) | 58 (98.31%) | 59 |
| Sum | 65 | 569 | 634 |

B
| Demographic |  | HC (n=114) | HIV (n=134) | Covid-Pos (n=327) | Covid-Neg (n=59) | p-value (all 4 cohorts) | p-value (CP/CN) |
| --- | --- | --- | --- | --- | --- | --- | --- |
| Race | Caucasian | 94 (82.46%) | 92 (68.66%) | 99 (30.28%) | 27 (45.76%) | <0.0001 | 0.1204 |
|  | African American | 17 (14.91%) | 36 (26.87%) | 223 (68.20%) | 32 (54.24%) |  |  |
|  | Asian | 3 (2.63%) | 5 (3.73%) | 2 (0.61%) | 0 (0.00%) |  |  |
|  | Other | 0 (0.00%) | 1 (0.75%) | 3 (0.92%) | 0 (0.00%) |  |  |
| Hispanic? (Y/N) | Yes | 2 (1.75%) | 8 (5.97%) | 0 (0.00%) | 0 (0.00%) | <0.0001 | >0.9999 |
|  | No | 112 (98.25%) | 126 (94.03%) | 327 (100.00%) | 59 (100.00%) |  |  |
| Sex | Male | 37 (32.46%) | 99 (73.88%) | 193 (59.02%) | 24 (40.68%) | <0.0001 | 0.4878 |
|  | Female | 77 (67.54%) | 32 (23.88%) | 134 (40.98%) | 35 (59.32%) |  |  |
|  | Other | 0 (0.00%) | 3 (2.24%) | 0 (0.00%) | 0 (0.00%) |  |  |
| Age (mean ± std dev) | - | 43.73 ± 16.94 | 46.99 ± 15.19 | 59.26 ± 16.52 | 47.02 ± 16.93 | <0.0001 | <0.0001 * |

**Table 2** compares demographic and clinical outcomes between SARS-CoV-2–positive (n = 327) and –negative (n = 59) patients. Mean BMI was similar in both groups (31.6 ± 9.4 vs. 30.6 ± 8.4; p = 0.51), but COVID-19–positive patients were far more likely to be hospitalized within 14 days (93.6% vs. 44.1%; p < 0.0001) and showed a non-significant trend toward higher ICU admission (51.3% vs. 38.5%; p = 0.23) and mechanical ventilation rates (56.1% vs. 40.0%; p = 0.055). Among ventilated patients, duration was markedly longer in the COVID-19 group (361.9 ± 444.7 vs. 54.8 ± 54.96 hours; p = 0.06). COVID-19 positivity was also associated with a longer time to discharge (12.3 ± 16.4 vs. 6.3 ± 9.8 days; p = 0.004) and higher mortality (24.5% vs. 10.2%; p = 0.0165), whereas no deaths in the SARS-CoV-2–negative cohort were attributed to COVID-19.

**Table 2.** Descriptive cohort table – Covid-Pos and -Neg subsets.

**Covid-Pos and -Neg patients<sup>1</sup>**
|  | **Covid-Pos (n=327)** | **Covid-Neg (n=59)** | **p-value** |
| --- | --- | --- | --- |
| **BMI** (mean ± std dev) | 31.58 ± 9.41 | 30.55 ± 8.35 | 0.5104 |
| **Hospitalization within 14 days** (%) | 306/327 (93.58%) | 26/59 (44.07%) | **<0.0001** * |
| **ICU admission<sup>2</sup>** (%) | 157/306 (51.31%) | 10/26 (38.46%) | 0.2265 |
| **Ventilation<sup>3</sup>** (%) | 88/157 (56.05%) | 4/10 (40.00%) | 0.0553 |
| **Ventilation Duration<sup>4</sup>** (mean ± std dev) | 361.9 ± 444.7 | 54.80 ± 54.96 | 0.0597 |
| **Discharge Status<sup>5</sup>** | 249 | 25 | - |
| **Time to Discharge<sup>6</sup>** (mean ± std dev) | 12.31 ± 16.40 (227) | 6.29 ± 9.78 (21) | **0.0042** * |
| **Time to Death<sup>6</sup>** (mean ± std dev) | 17.00 ± 16.16 (79) | 12.80 ± 6.94 (5) | 0.8508 |
| **Mortality Status: Alive** (%) | 247/327 (75.54%) | 53/59 (89.83%) | 0.0165 |
| **Death due to COVID-19** (%) | 63/80 (78.75%) | 0/6 (0.00%) | - |

<sup>1</sup>Some samples are missing hospitalization data, so proportions are from statistics available
<sup>2</sup>Of those hospitalized, proportion of patients admitted to ICU
<sup>3</sup>Of those admitted to ICU, proportion of patients ventilated
<sup>4</sup>Of those ventilated, ventilation duration statistics
<sup>5</sup>Only have records of "Yes"
<sup>6</sup>Time from hospitalization to either discharge or death - for death, includes decease in hospital and post-discharge, (number of data entries)
\*Significant under alpha = 0.05

To evaluate whether HPgV co-infection influences COVID-19 severity, we compared clinical outcomes (hospitalization rates, ICU admissions, and mortality) between COVID-19 patients with and without HPgV (**Table 3A-B, Additional File 1**). The overall number of positive patients was low, precluding robust statistical analyses.

**Table 3.**
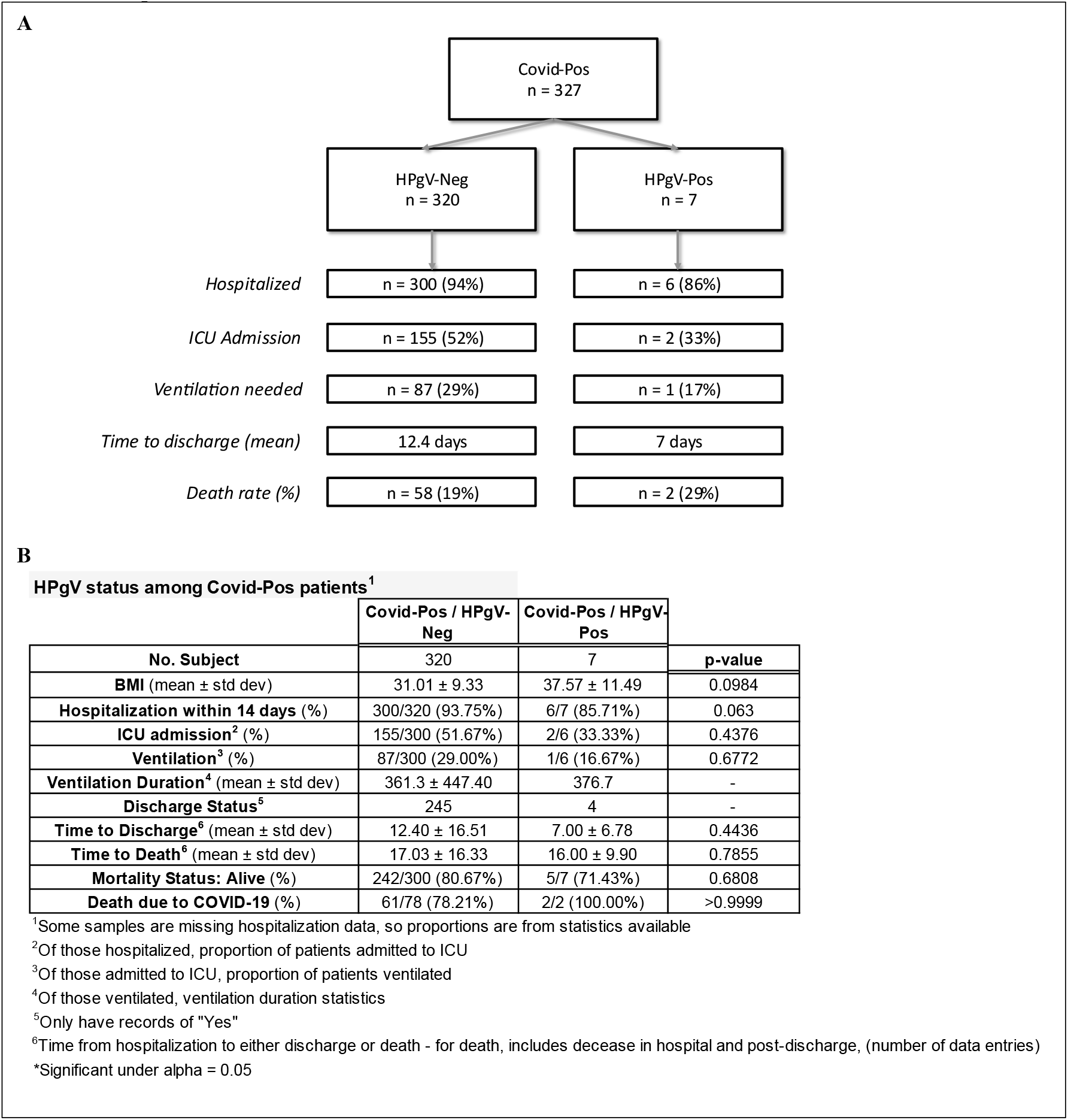
Descriptive cohort table – Covid-Pos subset.

|  | Covid-Pos / HPgV-Neg | Covid-Pos / HPgV-Pos | p-value |
| --- | --- | --- | --- |
| **No. Subject** | 320 | 7 |  |
| **BMI (mean ± std dev)** | 31.01 ± 9.33 | 37.57 ± 11.49 | 0.0984 |
| **Hospitalization within 14 days (%)** | 300/320 (93.75%) | 6/7 (85.71%) | 0.063 |
| **ICU admission<sup>2</sup> (%)** | 155/300 (51.67%) | 2/6 (33.33%) | 0.4376 |
| **Ventilation<sup>3</sup> (%)** | 87/300 (29.00%) | 1/6 (16.67%) | 0.6772 |
| **Ventilation Duration<sup>4</sup> (mean ± std dev)** | 361.3 ± 447.40 | 376.7 | - |
| **Discharge Status<sup>5</sup>** | 245 | 4 | - |
| **Time to Discharge<sup>6</sup> (mean ± std dev)** | 12.40 ± 16.51 | 7.00 ± 6.78 | 0.4436 |
| **Time to Death<sup>6</sup> (mean ± std dev)** | 17.03 ± 16.33 | 16.00 ± 9.90 | 0.7855 |
| **Mortality Status: Alive (%)** | 242/300 (80.67%) | 5/7 (71.43%) | 0.6808 |
| **Death due to COVID-19 (%)** | 61/78 (78.21%) | 2/2 (100.00%) | >0.9999 |

<sup>1</sup>Some samples are missing hospitalization data, so proportions are from statistics available<sup>2</sup>Of those hospitalized, proportion of patients admitted to ICU<sup>3</sup>Of those admitted to ICU, proportion of patients ventilated<sup>4</sup>Of those ventilated, ventilation duration statistics<sup>5</sup>Only have records of "Yes"<sup>6</sup>Time from hospitalization to either discharge or death - for death, includes decease in hospital and post-discharge, (number of data entries)
\*Significant under alpha = 0.05

Of the seven HPgV-positive COVID-19 patients, two died under exceptional circumstances. The first was a Caucasian man in his lower 80’s (range of 81-85 years old) with diabetes and hypertension who presented in extremis; given his advanced age and comorbidities, mechanical ventilation was intentionally withheld in favor of comfort measures. The second was an African American woman in her upper 30’s (range of 36-40 years old) with a BMI of 59, hypertension, peripheral vascular disease, and end-stage renal failure. Despite 376 hours of ICU-level mechanical ventilation, she succumbed to multi-organ complications.

Of note, however, the other five HPgV-positive patients experienced mild to moderate disease courses, despite having several risk factors for death following COVID infection [1 with diabetes (DM) and hypertension (HTN); 1 with DM, HTN, seizure disorder, paralysis and renal failure (RF); 1 with advanced liver disease; 1 with HTN, peripheral vascular disease, and RF; no data was available for 1 patient]. Furthermore, though not statistically significant, perhaps due to sample size, the median BMI for the HPgV+/COVID+ patients was 38 vs 31 for HPgV-/COVID+. None of these five HPgV+/COVID+ patients required ICU admission or ventilation, and one was managed entirely as an outpatient. Of the four hospitalized patients, one with a history of smoking had the longest stay (17 days), while the remaining three were discharged within 5 days of admission, notably shorter than the population median of 14 days for COVID-19 patients’ length of stay (16). Symptoms among HPgV-positive cases were consistent with typical COVID-19 presentations. Fever, cough, and diarrhea were the most frequently reported manifestations.

Comparison of COVID-19 outcomes in HPgV-positive (n=7) versus HPgV-negative (n=320) patients revealed consistent—but non–statistically significant—trends toward milder disease in the HPgV-positive group: the HPgV-positive group had lower rates of hospitalization (85.7% vs. 93.8%), ICU admission (33.3% vs. 51.7%), and mechanical ventilation (16.7% vs. 29.0%), and a shorter mean time to discharge (7.0 ± 6.8 vs. 12.4 ± 16.5 days). Although small numbers limit formal inference, these consistent directional differences raise the possibility that HPgV co-infection is associated with reduced COVID-19 severity (**Table 3B**).

## Discussion

This study provides a large-scale systematic evaluation of HPgV as an unmonitored viral co-infection in COVID-19, situated within the broader framework of viral interactions that may modify SARS-CoV-2 disease. HPgV prevalence among COVID-19 patients mirrored that of the general population, while the HIV cohort served as a robust positive control, reinforcing known associations between HPgV persistence and chronic viral infection. This unexpected elevation among healthcare workers highlights a potential occupational exposure risk. A recent study with healthcare workers, particularly those in pediatric and NICU units, showed immune alterations in response to repeated viral exposures that influence baseline T cell functionality and long-term immunity (13).

Although limited by small numbers, consistent trends toward lower hospitalization, ICU admission, ventilation, and shorter stays among HPgV-positive patients suggest that viral co-infections with immunomodulatory properties may influence SARS-CoV-2 pathogenesis.

In future work, a descriptive profiling of HPgV genotypes across different cohorts may provide insights into patterns of viral genetic variation in relation to co-infection. As well, it would be worth to analyze how HPgV co-infection shapes the SARS-CoV-2–specific antibody and memory B-cell repertoire. Indeed, similarly to the observations in measles infection (17), viral co-infection may alter immune profiles relevant to SARS-CoV-2 and other viral infections. Finally, further work should expand surveillance of healthcare workers across settings to delineate exposure and transmission patterns. It should be noted that our cross-sectional design and small HPgV-positive sample size preclude definitive conclusions. A larger longitudinal cohort with serial virologic and immunologic assessments will be critical to confirm whether HPgV truly is associated with milder COVID-19 outcomes and to evaluate its role in post-acute sequelae.

## Conclusion

In summary, HPgV co-infection in our COVID-19 cohort mirrored general population prevalence but showed consistent trends toward milder clinical courses. The high HPgV prevalence among healthcare-worker controls raises important questions about occupational exposure. While our small sample size precludes definitive conclusions, our findings support broader consideration of immunomodulatory viral co-infections in COVID-19 and motivate longitudinal studies to define their mechanistic and clinical significance.

## DECLARATION

### Author contributions

MG, PE *conceptualized* the study. MG, CCZ, PK, PE contributed to *methodology, investigation*, and *interpretation*. MG, CCZ, GG performed *formal analysis*. MG, CCZ, GG, PE *wrote the original draft*. PE provided *resources* and *funding*. All authors have read and approved the final manuscript. Criteria are as indicated by the contributor roles taxonomy.

### Additional information

#### Ethics Approval and Consent to Participate

The study was approved by the Washington University Institutional Review Board (IRB #202210110) with a determination of ‘non-human subject research.’ All plasma samples used in this study were de-identified, and no consent was required from participants.

### Data Availability

The datasets generated during and/or analyzed during the current study are available from the corresponding author on reasonable request.

## Supplemental Information

Additional information for Methods.

## Additional File

1. Table S1 - Clinical event and laboratory data of HPgV positive individuals with the Covid-Pos and -Neg cohort.

## References

1. Malveste Ito CR, Moreira ALE, Silva PAN da, Santos M de O, Santos APD, Rézio GS, et al. Viral Coinfection of Children Hospitalized with Severe Acute Respiratory Infections during COVID-19 Pandemic. Biomedicines. 2023 May 9;11(5):1402.

2. Shah MM, Patel K, Milucky J, Taylor CA, Reingold A, Armistead I, et al. Bacterial and viral infections among adults hospitalized with COVID-19, COVID-NET, 14 states, March 2020–April 2022. Influenza Other Respir Viruses. 2023 Mar 2;17(3):e13107.

3. Stapleton JT, Foung S, Muerhoff AS, Bukh J, Simmonds P. The GB viruses: a review and proposed classification of GBV-A, GBV-C (HGV), and GBV-D in genus Pegivirus within the family Flaviviridae. J Gen Virol. 2011 Feb;92(Pt 2):233–46.

4. Stapleton J. Human Pegivirus Type 1: A common human virus that is beneficial in immune-mediated disease. Frontiers in Immunology. 2022 May 30;13:17.

5. Chang CM, Stapleton JT, Klinzman D, McLinden JH, Purdue MP, Katki HA, et al. GBV-C INFECTION AND RISK OF NHL AMONG U.S. ADULTS. Cancer Res. 2014 Oct 1;74(19):5553–60.

6. Krajden M, Yu A, Braybrook H, Lai AS, Mak A, Chow R, et al. GBV-C/hepatitis G virus infection and non-Hodgkin lymphoma: a case control study. International Journal of Cancer. 2010;126(12):2885–92.

7. Fama A, Xiang J, Link BK, Allmer C, Klinzman D, Feldman AL, et al. Human Pegivirus infection and lymphoma risk and prognosis: A North American study. Br J Haematol. 2018 Sept;182(5):644–53.

8. Fama A, Larson MC, Link BK, Habermann TM, Feldman AL, Call TG, et al. Human Pegivirus Infection and Lymphoma Risk: A Systematic Review and Meta-analysis. Clin Infect Dis. 2020 Aug 22;71(5):1221–8.

9. Wu Zhang X, Leng Yap Y. Structural similarity between HIV-1 gp41 and SARS-CoV S2 proteins suggests an analogous membrane fusion mechanism. Theochem. 2004 May;677(1):73–6.

10. Fantini J, Chahinian H, Yahi N. Convergent Evolution Dynamics of SARS-CoV-2 and HIV Surface Envelope Glycoproteins Driven by Host Cell Surface Receptors and Lipid Rafts: Lessons for the Future. Int J Mol Sci. 2023 Jan 18;24(3):1923.

11. Mannar D, Leopold K, Subramaniam S. Glycan reactive anti-HIV-1 antibodies bind the SARS-CoV-2 spike protein but do not block viral entry. Sci Rep. 2021 June 14;11(1):12448.

12. Perween R, PraveenKumar M, Shrivastava T, Parray HA, Singh V, Singh S, et al. The SARS CoV-2 spike directed non-neutralizing polyclonal antibodies cross-react with Human immunodeficiency virus (HIV-1) gp41. Int Immunopharmacol. 2021 Dec;101(Pt B):108187.

13. Elias G, Souquette A, Heynderickx S, De Meester I, Jansens H, Beutels P, et al. Altered CD4+ T cell immunity in nurses occupationally exposed to viral pathogens. Clin Exp Immunol. 2018 Nov;194(2):192–204.

14. Tissue Procurement Core (TCP) WashU. Washington University’s COVID-19 Biorepository. 2024 [Internet]. Available from: https://icts.wustl.edu/icts-highlights-impacts-made-from-covid-19-biorepository/

15. Rydze RT, Bhattarai N, Stapleton JT. GB virus C infection is associated with a reduced rate of reactivation of latent HIV and protection against activation-induced T-cell death. Antivir Ther. 2012;17(7):1271–9.

16. Alimohamadi Y, Yekta EM, Sepandi M, Sharafoddin M, Arshadi M, Hesari E. Hospital length of stay for COVID-19 patients: a systematic review and meta-analysis. Multidiscip Respir Med. 2022 Jan 12;17(1):856.

17. Mina MJ, Kula T, Leng Y, Li M, de Vries RD, Knip M, et al. Measles virus infection diminishes preexisting antibodies that offer protection from other pathogens. Science. 2019 Nov 1;366(6465):599–606.

