## Supplemental Information for "Viral Co-infection in COVID-19: Prevalence and Clinical Associations of Human Pegivirus"

**SUPPLEMENTAL INFORMATION - SI**

To detect HPgV viral nucleic acid via RT-qPCR, we follow a precise protocol. We start by extracting total nucleic acids from 100 µL of plasma using the Zymo Quick DNA/RNA Viral Kit and eluting them in 30 µL of DNase/RNase-free water. We then assess the concentration and quality ratio using a Nanodrop and store the samples at -80°C until further analysis. For RNA quantification, we employ quantitative one-step real-time RT-PCR (RT-qPCR) with the SuperScript® III Platinum® One-Step qRT-PCR Kit, following the manufacturer's protocol. The specific primers used for HPgV RNA amplification are as follows: Forward Primer 5′- GGCGACCGGCCAAAA -3′ and Reverse Primer 5′-GTAGCCACTATAGGTGGGTCTTAAG -3′. We also use the TaqMan/MGB probe: 5′-AGGGTTGGTAGGTCGTAAATCCCGGTCA-3′, as previously described (15). We conduct the RT-qPCR on a QuantStudio3 instrument, with the following running conditions: 50°C for 20 min, followed by 95°C for 2 min, and then 40 cycles at 95°C for 15 sec and 58°C for 1 min. To ensure the reliability of our results, we maintain linear regression analysis of standard curves with an R2 > 0.98 and ensure that the variation between cycle threshold (Ct) values between experiments remains within 1.5 Ct. Each assay reaction employs 11 µL of plasma nucleic acid, and we calibrate our assays using an in vitro transcribed standard. The plasmid used for transcribing the RNA standard spans from the 5' end of the genome through nt 850, based on the sequence of the Hepatitis G virus strain Iowan, complete genome (AF121950.1 GI:4884678) (15). In brief, the plasmid is linearized, and transcripts are generated, quantifying them through absorbance and terminal dilution using the primer/probe set corresponding to this portion of the genome. We obtained purified RNA standards (10^8 transcripts/mL) from our Dr. Stapleton. Following qRT-PCR, all samples with a Ct value of less than 35, i.e. classified as HPgV positive, and a selected number of HPgV negative samples were subjected to gel electrophoresis to validate the HPgV status results (representative image shown below).

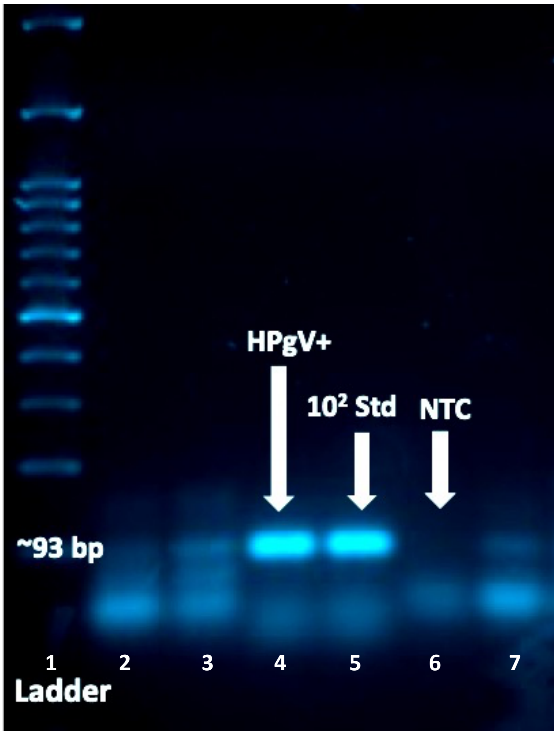


*Lanes 1:1kb ladder, 2:negative sample, 3:positive sample (non-specific band visible), 4:positive control, 5:RNA standard 10^2, 6:negative control. Excess primers are visible as the lowest band in each lane.*
