## Additional File 1 for "Viral Co-infection in COVID-19: Prevalence and Clinical Associations of Human Pegivirus"

| Participant ID | WU350-086 | WU350-093 | WU350-128 | WU350-329 | WU350-361 | WU350-445 | WU350-490 |
|---|---|---|---|---|---|---|---|
| COVID | Positive | Positive | Positive | Positive | Positive | Positive | Positive |
| days since first biospecimen | 0 | 0 | 0 | 0 | 0 | 0 | 0 |
| Ct value 1 | 30.9 | 21.2 | 33.7 | 24.988 | 31.5 | 19.4 | 23.548 |
| HPgV: Positive / Negative | Positive | Positive | Positive | Positive | Positive | Positive | Positive |
| Age Range at first biospecimen sample (years) | 41-45 | 81-85 | 26-30 | 31-35 | 51-55 | 26-30 | 36-40 |
| Sex | Female | Male | Male | Male | Male | Female | Female |
| Epic BMI: Closest to biospecimen sample 1 | 42.8 | 28.3 | 43.9 | 29.2 | 35.6 | 27.2 | 59.3 |
| Race | African American | White | African American | African American | African American | White | African American |
| Epic: Index Hospitalization (within 14 days of biospecimen #1) | Yes | Yes | Yes | No | Yes | Yes | Yes |
| Epic: ICU admission during index hospital visit | No | Yes | No | *nap* | No | No | Yes |
| Subject on ventilation during index hospital visit | No | No | No | *nap* | No | No | Yes |
| Total Ventilation Duration during index hospital visit (hours) | *nap* | *nap* | *nap* | *nap* | *nap* | *nap* | 376.7 |
| Ventilation unrelated to respiratory failure? | *nap* | *nap* | *nap* | *nap* | *nap* | *nap* | *nap* |
| Extubated and alive during index hospital visit | *nap* | *nap* | *nap* | *nap* | *nap* | *nap* | No |
| Epic: Discharged | Yes | *nap* | Yes | *nap* | Yes | Yes | *nap* |
| Epic: Readmitted to hospital | Yes | No | Yes | *nap* | No | Yes | No |
| Epic: Mortality status | Alive | Decease d | Alive | Alive | Alive | Alive | Deceased |
| Epic: Died during index visit? | *nap* | Yes | *nap* | *nap* | *nap* | *nap* | Yes |
| Death due to COVID-19? | *nap* | Yes | *nap* | *nap* | *nap* | *nap* | Yes |
| Epic: Time from symptom onset to index hospitalization (days) | 14 | 2 | 2 | *nap* | 3 | 50 | 3 |
| Epic: Time from index hospitalization to discharge or death during hospitalization (days) | 4 | 9 | 2 | *nap* | 5 | 17 | 23 |
| Epic: Time from symptom onset to death (days) | *nap* | 11 | *nap* | *nap* | *nap* | *nap* | 26 |
| Time from discharge from index hospitalization to re-admission to hospital (days) | 274 | *nap* | 1 | *nap* | *nap* | 105 | *nap* |
| Epic: Time from symptom onset to ICU (days) | *nap* | 3 | *nap* | *nap* | *nap* | *nap* | 4 |
| Other coronavirus positive: | *n/a* | No | No | *n/a* | *n/a* | *n/a* | No |
| Is this patient homeless? | No | No | No | No | No | No | No |

| | | | | | | | |
|---|---|---|---|---|---|---|---|
| Has the patient ever used tobacco products? | No | Yes | No | Yes | No | Yes | No |
| If patient has ever used tobacco, has the patient smoked tobacco? | No | Yes | No | Yes | No | Yes | No |
| If yes to smoked tobacco, is the patient a current smoker? | *nap* | No | *nap* | Yes | *nap* | No | *nap* |
| Has the patient ever used vaping products? | No | No | No | No | No | No | No |
| If yes, does the patient currently use vaping products? | *nap* | *nap* | *nap* | *nap* | *nap* | *nap* | *nap* |
| Has this patient had contact with a person with a known diagnosis of SARS-CoV-2 in the last 14 days? | No | No | No | No | Yes | No | No |
| Has this patient had contact with a person with a respiratory tract illness in the last 14 days? | No | No | No | No | Yes | No | No |
| Has the patient traveled in the last 90 days? | No | No | No | No | No | No | No |
| Domestic travel in past 90 days (if answer yes to any travel in past 90 days) | No | No | No | No | No | No | No |
| States traveled to in past 90 days | *nap* | *nap* | *nap* | *nap* | *nap* | *nap* | *nap* |
| International travel in past 90 days (if answer yes to any travel in past 90 days) | No | No | No | No | No | No | No |
| International locations traveled to in past 90 days | *nap* | *nap* | *nap* | *nap* | *nap* | *nap* | *nap* |
| Symptom: Fever | Yes | Yes | No | No | No | Yes | Yes |
| Symptom: Chills | Yes | No | No | No | No | Yes | No |
| Symptom: Conjunctival congestion | No | No | No | No | No | No | No |
| Symptom: Nasal congestion | No | No | No | No | No | No | No |
| Symptom: Headaches | No | No | No | No | No | No | No |
| Symptom: Cough | No | Yes | Yes | Yes | No | Yes | No |
| Symptom: Sputum production | No | No | No | No | No | Yes | No |
| Symptom: Sore throat | No | Yes | No | No | No | No | No |
| Symptom: Shortness of breath | No | No | Yes | No | No | No | Yes |
| Symptom: Nausea or vomiting | Yes | No | No | No | No | Yes | No |
| Symptom: Diarrhea | Yes | No | Yes | No | Yes | Yes | No |
| Symptom: Myalgia | Yes | No | No | No | Yes | Yes | No |
| Symptom: Fatigue | No | No | No | Yes | No | Yes | Yes |
| Symptom: Rash | No | No | No | No | No | No | No |
| Symptom: Lymphadenopathy | No | No | No | No | No | No | No |
| Symptom: Confusion | No | No | No | No | No | No | No |
| Symptom: Other | No | No | No | Yes | No | Yes | No |
| Specify other symptoms: | *nap* | *nap* | *nap* | loss of taste and smell | *nap* | loss of smell and taste | *nap* |
| Asymptomatic | No | No | No | No | No | No | No |

| | | | | | | | |
|---|---|---|---|---|---|---|---|
| **Comorbidities_1yr** | | | | | | | |
| Acquired immune deficiency syndrome | 0 | 0 | *n/a* | 0 | 0 | 0 | 0 |
| Alcohol abuse | 0 | 0 | *n/a* | 0 | 0 | 0 | 0 |
| Deficiency anemias | 1 | 0 | *n/a* | 0 | 0 | 0 | 0 |
| Arthropathies | 0 | 0 | *n/a* | 0 | 0 | 0 | 0 |
| Chronic blood loss anemia | 0 | 0 | *n/a* | 0 | 0 | 0 | 0 |
| Leukemia | 0 | 0 | *n/a* | 0 | 0 | 0 | 0 |
| Lymphoma | 0 | 0 | *n/a* | 0 | 0 | 0 | 0 |
| Cancer: Metastic OR solid tumor w/o metastisis (in situ) OR solid tumor w/o metastisis (malignant) | 0 | 0 | *n/a* | 0 | 0 | 0 | 0 |
| Cerebrovascular disease | 0 | 0 | *n/a* | 0 | 1 | 0 | 0 |
| Congestive heart failure | 0 | 0 | *n/a* | 0 | 1 | 0 | 0 |
| Coagulopathy | 0 | 0 | *n/a* | 0 | 0 | 0 | 0 |
| Dementia | 0 | 0 | *n/a* | 0 | 0 | 0 | 0 |
| Depression | 0 | 0 | *n/a* | 0 | 0 | 0 | 0 |
| Diabetes with or without chronic complications | 1 | 1 | *n/a* | 0 | 1 | 0 | 0 |
| Drug abuse | 0 | 0 | *n/a* | 0 | 0 | 0 | 0 |
| Hypertension, complicated or uncomplicated | 1 | 1 | *n/a* | 0 | 1 | 0 | 1 |
| Liver disease | 0 | 0 | *n/a* | 0 | 0 | 1 | 0 |
| Chronic pulmonary disease | 0 | 0 | *n/a* | 0 | 0 | 0 | 0 |
| Neurological disorders affecting movement | 0 | 0 | *n/a* | 0 | 0 | 0 | 0 |
| Other neurological disorders | 0 | 0 | *n/a* | 0 | 1 | 0 | 0 |
| Seizures and epilepsy | 0 | 0 | *n/a* | 0 | 1 | 0 | 0 |
| Obesity | 0 | 0 | *n/a* | 0 | 0 | 0 | 0 |
| Paralysis | 0 | 0 | *n/a* | 0 | 1 | 0 | 0 |
| Peripheral vascular disease | 0 | 0 | *n/a* | 0 | 0 | 0 | 1 |
| Psychoses | 0 | 0 | *n/a* | 1 | 0 | 0 | 0 |
| Pulmonary circulation disease | 0 | 0 | *n/a* | 0 | 0 | 0 | 0 |
| Renal failure | 0 | 0 | *n/a* | 0 | 1 | 0 | 1 |
| Hypothyroidism | 0 | 0 | *n/a* | 0 | 0 | 0 | 0 |
| Other thyroid disorders | 0 | 0 | *n/a* | 0 | 0 | 0 | 0 |
| Peptic ulcer with bleeding | 0 | 0 | *n/a* | 0 | 0 | 0 | 0 |
| Valvular disease | 0 | 0 | *n/a* | 0 | 0 | 0 | 0 |
| Weight loss | 0 | 0 | *n/a* | 0 | 0 | 0 | 0 |
| **Labs 24hrs** | | | | | | | |
| ALANINE AMINOTRANSFERASE (Units/L) | 41 | 21 | 182 | 37 | *n/a* | 50 | 73 |
| ALBUMIN (g/dL) | 3.3 | 3 | 4 | 4.1 | 2.9 | 2.4 | 2.8 |
| ALKALINE PHOSPHATASE (Units/L) | 47 | 53 | 91 | 64 | *n/a* | 257 | 62 |
| ANION GAP (mmol/L) | 10 | 9 | 14 | 8 | 13 | 7 | 15 |

| | | | | | | | |
|---|---|---|---|---|---|---|---|
| ASPARTATE AMINOTRANSFERASE (Units/L) | 77 | 13 | 264 | 37 | *n/a* | 67 | 58 |
| BASOPHIL ABSOLUTE (K/cumm) | 0 | 0 | 0 | 0 | 0 | 0.2 | 0 |
| BASOPHIL PERCENT (%) | 0 | 0.2 | 0.2 | 0.2 | 0.2 | 0.5 | 0.1 |
| BILIRUBIN, TOTAL (mg/dL) | 0.4 | 0.7 | 1.6 | 0.4 | *n/a* | 1.1 | 0.3 |
| CALCIUM, TOTAL (mg/dL) | 8.5 | 8.7 | 8.5 | 9.2 | 8.5 | 7 | 8.5 |
| CHLORIDE (mmol/L) | 99 | 107 | 93 | 102 | 108 | 96 | 99 |
| CO2, TOTAL (mmol/L) | 26 | 23 | 18 | 27 | 18 | 25 | 27 |
| CREATININE (mg/dL) | 1.35 | 2.14 | 1.08 | 1.33 | 3.74 | 0.71 | 5.61 |
| EOSINOPHIL ABSOLUTE (K/cumm) | 0 | 0 | 0 | 0 | 0 | 0 | 0 |
| EOSINOPHIL PERCENT (%) | 0 | 0.2 | 0 | 0.5 | 0.4 | 0.1 | 0 |
| GLUCOSE (mg/dL) | 139 | 218 | 132 | 88 | 149 | 123 | 106 |
| HEMATOCRIT (%) | 39.7 | 23.1 | 42.7 | 43.8 | 21.8 | 30.9 | 27.2 |
| HEMOGLOBIN (g/dL) | 13.3 | 7.7 | 15.5 | 14.7 | 7 | 11.7 | 8.9 |
| IMMATURE GRANULOCYTE ABSOLUTE (K/cumm) | 0 | 0 | 0.1 | 0 | 0 | 1 | 0.1 |
| IMMATURE GRANULOCYTE PERCENT (%) | 0.4 | 0.4 | 0.8 | 0.2 | 0.2 | 2.5 | 0.7 |
| LYMPHOCYTE ABSOLUTE (K/cumm) | 1.8 | 0.2 | 0.6 | 1.6 | 1.7 | 0.4 | 0.9 |
| LYMPHOCYTE PERCENT (%) | 23.8 | 2.9 | 9.7 | 36.8 | 36.7 | 1 | 6.8 |
| MEAN CELLULAR HEMOGLOBIN (pg) | 28.1 | 27.6 | 28.5 | 28.7 | 29.3 | 35.8 | 30 |
| MEAN CELLULAR HEMOGLOBIN CONCENTRATION (g/dL) | 33.5 | 33.3 | 36.3 | 33.6 | 32.1 | 37.9 | 32.7 |
| MEAN CELLULAR VOLUME (fL) | 83.8 | 82.8 | 78.5 | 85.4 | 91.2 | 94.5 | 91.6 |
| MEAN PLATELET VOLUME (fL) | 12.8 | 10 | 10.9 | 11.9 | 11 | 12.6 | 10.2 |
| MONOCYTE ABSOLUTE (K/cumm) | 0.3 | 0.4 | 0.5 | 0.4 | 0.5 | 3.1 | 0.8 |
| MONOCYTE PERCENT (%) | 4 | 7.1 | 7.7 | 8.4 | 10 | 8.1 | 6.4 |
| NEUTROPHIL ABSOLUTE (K/cumm) | 5.4 | 4.5 | 5.1 | 2.3 | 2.4 | 34 | 11 |
| NEUTROPHIL PERCENT (%) | 71.8 | 89.2 | 81.6 | 53.9 | 52.5 | 87.8 | 86 |
| NUCLEATED RBC ABS, AUTO (K/cumm) | 0 | 0 | 0 | 0 | 0 | 0.02 | 0 |
| PLATELET (K/cumm) | 194 | 139 | 222 | 165 | 211 | 154 | 211 |
| POTASSIUM, PLASMA (mmol/L) | 3.5 | 4.2 | 4.5 | 4.7 | 4 | 3.3 | 3.3 |
| PROTEIN, TOTAL, PLASMA (g/dL) | 7.9 | 5.4 | 8.2 | 7.3 | *n/a* | 6.6 | 5.9 |
| RED BLOOD CELL (M/cumm) | 4.74 | 2.79 | 5.44 | 5.13 | 2.39 | 3.27 | 2.97 |
| RED CELL DISTRIBUTION WIDTH CV (%) | 13.2 | 15.1 | 13.5 | 13 | 11.8 | 17.9 | 14.1 |

| | | | | | | | |
|---|---|---|---|---|---|---|---|
| RED CELL DISTRIBUTION WIDTH SD (fL) | 40.1 | 45.5 | 38.4 | 40.2 | 39.8 | 62.2 | 46.9 |
| SODIUM (mmol/L) | 135 | 139 | 125 | 137 | 139 | 128 | 141 |
| UREA NITROGEN SERUM (mg/dL) | 14 | 48 | 17 | 9 | 75 | 17 | 120 |
| WHITE BLOOD CELL COUNT (K/cumm) | 7.5 | 5.1 | 6.2 | 4.3 | 4.5 | 38.7 | 12.8 |
| MAGNESIUM (mg/dL) | 1.5 | 2.1 | 2.4 | *n/a* | 2.2 | 1 | 1.9 |
| D-DIMER (ng/mL FEU) | 1199 | 1406 | 1295 | *n/a* | 695 | *n/a* | 13243 |
| FERRITIN (ng/mL) | *n/a* | 2891 | 1365 | *n/a* | 2492 | *n/a* | *n/a* |
| INTERNATIONAL NORMALIZED RATIO () | *n/a* | *n/a* | 1.5 | *n/a* | *n/a* | 1.5 | 1 |
| CREATINE KINASE, TOTAL (Units/L) | *n/a* | 60 | 339 | *n/a* | *n/a* | *n/a* | *n/a* |
| C REACTIVE PROTEIN (mg/L) | 138.6 | 158.3 | 78.1 | *n/a* | 65 | 246.1 | 57.1 |
| LACTATE DEHYDROGENASE, TOTAL (Units/L) | *n/a* | *n/a* | 705 | *n/a* | 299 | *n/a* | *n/a* |
| ERYTHROCYTE SEDIMENTATION RATE (mm/hr) | 74 | 70 | 36 | *n/a* | *n/a* | 12 | *n/a* |

*NOTES:*
*nap = not applicable*
*n/a = not available*
